# A head-to-head comparison of plasma biomarkers to detect biologically defined Alzheimer in a memory clinic

**DOI:** 10.1101/2024.10.26.24316176

**Authors:** Federica Anastasi, Aida Fernández-Lebrero, Nicholas J. Ashton, Paula Ortiz-Romero, Javier Torres-Torronteras, Armand González-Escalante, Marta Milà-Alomà, José Contador, Greta García-Escobar, Rosa María Manero-Borràs, Irene Navalpotro-Gómez, Aparna Sahajan, Qinyu Hao, Bingqing Zhang, Andreas Jeromin, Nathalie Le Bastard, Alicia Nadal, Tahmine Mousavi, Gwendlyn Kollmorgen, Margherita Carboni, Oriol Grau-Rivera, Henrik Zetterberg, Marta del Campo, Kaj Blennow, Albert Puig-Pijoan, Marc Suárez-Calvet

## Abstract

**INTRODUCTION:** Blood-based biomarkers for Alzheimer’s disease (AD) have been widely studied, but direct comparisons of several biomarkers in clinical settings remain limited.

**METHODS:** In this cross-sectional study, plasma biomarkers from 197 participants in the BIODEGMAR cohort at Hospital del Mar (Barcelona) were analysed. Participants were classified based on AD cerebrospinal fluid (CSF) core biomarkers. We assessed the ability of plasma p-tau181, p-tau217, p-tau231, t-tau, and Aβ42/40 to classify Aβ status.

**RESULTS:** Plasma p-tau biomarkers had a greater diagnostic performance and larger effect sizes compared to t-tau and Aβ42/40 assays in detecting biologically defined AD. Among them, plasma p-tau217 consistently outperformed the others, demonstrating superior AUC. Furthermore, p-tau217 showed the strongest correlation between plasma and CSF levels, underscoring its potential as a reliable surrogate for CSF biomarkers.

**DISCUSSION:** Several plasma biomarkers, targeting different epitopes and using different platforms, demonstrated high performance in distinguishing biologically defined AD in a memory clinic setting.

## 1. BACKGROUND

While numerous studies have explored the diagnostic performance of blood-based biomarkers for detecting Alzheimer’s disease (AD),^1–10^ relatively few have conducted a comprehensive head-to-head comparison of multiple biomarkers in patients presenting at memory clinics,^11–16^ where patients often exhibit a broad spectrum of symptoms. Blood-based biomarkers offer a less invasive and accessible diagnostic tool, facilitating the etiological diagnosis of AD. Among these, assays targeting phosphorylated forms of tau, particularly at threonine 217 (T217), have demonstrated the highest diagnostic performance.^6,9–13^ In fact, p-tau217 assays have shown a comparable ability to detect AD pathology, as assessed by amyloid and tau PET scans, to that of core AD biomarkers in cerebrospinal fluid (CSF).^4^ Assays targeting phosphorylation at threonine 181 (T181)^5–7^ and 231 (T231)^8,17,18^ have also yielded promising results. Additionally, some assays targeting amyloid-β (Aβ) in plasma have shown favourable clinical performance.^1–3,15^ While immunoprecipitation mass spectrometry (IP-MS)-based assays generally achieve higher accuracy,^4,11,15^ certain immunoassays are also proving to be highly accurate for AD detection and are better suited for implementation in routine clinical laboratories.^9–12,19^

In a previous study, we compared the diagnostic performance of nine plasma assays targeting tau in a memory clinic cohort from a public hospital, using AD CSF core biomarkers as the biological definition of AD.^12^ Given the rapid development of new plasma biomarkers, this study incorporates additional plasma biomarker immunoassays and conducts a direct comparison in a real-world memory clinic population. Our research now encompasses results from up to 22 different plasma biomarker assays in a prospective clinical cohort, providing clinically relevant insights into how these biomarkers perform in routine practice.

## 2. METHODS

### 2.1 Participants

This cross-sectional study enrolled 197 participants from the BIODEGMAR cohort, an observational longitudinal study at the Cognition and Behaviour Unit of Hospital del Mar (Barcelona, Spain).^20^ The BIODEGMAR cohort comprises patients presenting with cognitive and/or behavioural symptoms who are derived from primary care to Hospital del Mar, a public university hospital, for further investigation. A lumbar puncture is performed as part of the routine clinical investigations (see supplementary material for more information).

The inclusion criteria of the BIODEGMAR cohort are: (i) evaluation at the Cognition and Behaviour Unit and inclusion in the DEGMAR register; (ii) signed informed consent; and (iii) having one of the following clinical diagnoses: subjective cognitive decline (SCD); mild cognitive impairment syndrome (MCI); AD dementia; behavioural variant frontotemporal dementia (bvFTD); progressive aphasia or primary progressive aphasia (logopenic, non-fluent and semantic variants, PA); Lewy body dementia (LBD); corticobasal syndrome (CBS); progressive supranuclear palsy syndrome (PSP-S) and vascular cognitive impairment and dementia (VCID). Individuals with other causes of dementia or unspecified clinical diagnoses were categorized as “other”. The exclusion criteria are: (i) age ≥80 years; (ii) contraindication for lumbar puncture; or (iii) disagreement with study procedures. The clinical criteria used for each diagnosis are indicated in Table S1.

Participants in the BIODEGMAR cohort included in the present study were recruited prospectively from April 2017 to July 2020, and fulfilled the following criteria: (i) had a Global Deterioration Score (GDS)>1^21^, (ii) Aβ42 and p-tau181 in CSF were available, and (iii) had at least one plasma biomarker available.

### 2.2 Plasma and CSF collection and storage. Biomarkers’ measurements

This study evaluated new nine p-tau and one tau assays: Lumipulse p-tau181, Meso Scale Discovery (MSD^®^) S-PLEX^®^ p-tau181, NULISA p-tau181, Roche Diagnostics using the NeuroToolKit p-tau181 (Roche p-tau181), ALZpath Simoa p-tau217, Lumipulse p-tau217, MSD S-PLEX p-tau217, NULISA p-tau217, NULISA p-tau231, along with the plasma t-tau assay NULISA MAPT. Additionally, three Aβ42/Aβ40 plasma assays were assessed: Lumipulse Aβ42/Aβ40, NULISA Aβ42/Aβ40, and Roche Diagnostics using the NeuroToolKit Aβ42/Aβ40 (Roche Aβ42/Aβ40). Detailed descriptions of these immunoassays are provided in the Methods section of the supporting information. The epitope binding regions for the assays are indicated in Table S2. NULISA assays were from a multiplex CNS Disease Panel NULISAseq assay.^22^

In addition to plasma measurements, corresponding CSF measurements were obtained for Lumipulse p-tau181, MSD p-tau181, NULISA p-tau181, Roche p-tau181, Lumipulse p-tau217, MSD p-tau217, NULISA p-tau217, NULISA p-tau231, and NULISA MAPT, from the same patients.

Blood and CSF samples were collected simultaneously under fasting conditions (at least 8 hours). Lumbar puncture was performed in the intervertebral space L3/L4, L4/L5 or L5/S1 using a standard needle, between 8 and 11 am. Participants had fasted for at least 8h. CSF was collected into a 10 ml sterile polypropylene sterile tube (Sarstedt, Nümbrecht, Germany; cat. no. 62.610.201). Tubes were gently inverted 5 – 10 times and centrifuged at 2000*g* for 10 minutes at 4°C and aliquoted in volumes of 1.8 ml into sterile polypropylene tubes (1.8 ml cryotube Thermo Scientific™ Nunc™; Thermo Fisher Scientific, Waltham, MA, USA; cat. No 377267), and immediately frozen at −80°C. Whole blood was drawn with a 20g or 21g needle gauge into 10 ml ethylenediaminetetraacetic acid (EDTA) tubes (BD Vacutainer 10 ml; K2EDTA; cat. no. 367525). Tubes were gently inverted 5 – 10 times and centrifuged at 2000*g* for 10 minutes at (4°C). The supernatant was aliquoted in volumes of 1.8 ml into sterile polypropylene tubes (1.8 ml cryotube Thermo Scientific™ Nunc™; Thermo Fisher Scientific, Waltham, MA, USA; cat. No 377267), and immediately frozen at −80°C.

AD CSF core biomarkers (Aβ42, Aβ40, p-tau181, and t-tau) were measured using the LUMIPULSE G600II (Fujirebio) at the Laboratori de Referència de Catalunya (Barcelona, Spain). Patients were classified as having an AD CSF profile if the CSF Aβ42/p-tau181 ratio was <10.25.^12,20^ In a subset of patients, these biomarkers were also measured using the Elecsys^®^ Phospho-Tau(181P) and β-Amyloid(1-42) CSF immunoassays (Roche Diagnostics International Ltd, Rotkreuz, Switzerland) at the Clinical Neurochemistry Laboratory at the University of Gothenburg (Sweden). Patients were classified as having an AD CSF profile if the CSF p-tau181/Aβ42 ratio was >0.029.^23^

Biomarker measurements were performed on aliquots that had undergone one freeze-thaw cycle, except for the ALZpath Simoa p-tau217 assay, which was measured after two freeze-thaw cycles. The assays were conducted at various specialized laboratories: Lumipulse p-tau181, ALZpath Simoa p-tau217, Lumipulse p-tau217, Lumipulse Aβ40 and Lumipulse Aβ42 assays at the Biomarkers Laboratory at BBRC (Barcelona, Spain); Roche p-tau181, Aβ40 and Aβ42 assays at the Clinical Neurochemistry Laboratory at the University of Gothenburg (Sweden); MSD p-tau181 and MSD p-tau217 assays at the MSD Bioanalytical Laboratory (BAL) (Gaithersburg, Maryland, USA); and NULISA assays at Alamar headquarters (Fremont, California, USA). Samples were shipped on dry ice. All technicians performing the measurements were blinded to clinical data, and data unblinding and analysis were conducted independently by study coordinators at BBRC. To ensure a direct comparison, all plasma and CSF biomarker measurements were taken from the same patients, with blood and CSF samples collected and processed simultaneously. However, some patients had missing biomarker data. To address this and maintain comparability, we conducted sensitivity analyses that included only participants with complete biomarker data. In this latter analysis, to ensure a comprehensive comparison across a wide range of plasma biomarkers, we also included the following previously measured biomarkers^12^: ADx p-tau181, Lilly p-tau181, Quanterix p-tau181, University of Gothenburg (UGot) in-house assay p-tau181, Lilly p-tau217, Janssen p-tau217, ADx p-tau231, UGot in-house assay p-tau231, and Lilly t-tau. A summary of the methods is available in the supplementary material, and Table S2 presents the key assay characteristics.

### 2.3 Statistical analyses

Differences in participant characteristics between the AD CSF profile group and the non-AD CSF profile groups were assessed using either a Mann-Whitney *U* test or Pearson’s χ² test, as appropriate. Plasma and CSF biomarkers did not meet the assumption of normally distributed residuals; therefore, non-parametric tests were applied. Outliers were not removed from the analysis.

Biomarker levels in the AD CSF profile group and the non-AD CSF profile group were presented as medians and interquartile ranges (IQRs). Differences between these two groups were evaluated using the Mann-Whitney *U* test. Effect sizes for the comparisons (r) were calculated by dividing the absolute standardized test statistic (Z) by the square root of the total number of individuals (*n*) or by indicating the percentage change of the median.

Receiver operating characteristic (ROC) analyses were conducted to assess the ability of biomarkers to discriminate between AD CSF profile from the non-AD CSF profile. Areas under the curve (AUC) and their 95% confidence intervals (CIs) were calculated using bootstrapping (n=2000 resamples with replacement). Optimal cutoffs for each tau biomarker were determined using the highest Youden’s index (sensitivity + specificity - 1) or setting the sensitivity at 90% or the specificity at 90%. Spearman rank-order correlation was used to assess correlations between biomarkers.

Sensitivity analyses were performed on the subgroup of patients who had all plasma biomarkers available. Additional sensitivity analyses were conducted in participants with syndromic diagnoses of SCD, MCI, and dementia, as well as in those at predementia stages (SCD and MCI).

All tests were two-tailed, with a significance level set at α = 0.05. Statistical analyses were performed using R version 4.4.0.

## 3. RESULTS

### 3.1 Patient characteristics

We included a total of 197 patients (Table 1), who had the following syndromic diagnosis: 18 SCD, 79 MCI, 62 AD dementia, 5 bvFTD, 12 PA, 4 LBD, 4 CBS, 3 PSP-S, 6 VCID and 4 classified as other dementia syndromes (Table S1).

All patients, irrespective of the clinical diagnosis, were classified biologically based on their AD CSF core biomarker profile (namely the Lumipulse CSF Aβ42/p-tau181 ratio) into AD (n=124) and non-AD (n=73) CSF profiles. Table 1 presents the main demographic and clinical characteristics of the samples. Participants with AD CSF profile were older and performed poorer on MMSE. A higher percentage of *APOE* ε4 carriers was found in the AD CSF profile group. Regarding sex differences, a higher percentage of women in the AD CSF profile group was observed, which did not reach statistical significance.

### 3.2 Plasma biomarker levels in biologically defined AD

Plasma biomarker levels were compared between patients with an AD and non-AD CSF profile (Table 2 and Figure 1). All plasma tau biomarkers were statistically significantly increased and all plasma Aβ42/40 biomarkers were significantly decreased in the AD CSF group compared to the non-AD CSF group. Nevertheless, the effect size of these differences varied between plasma biomarkers. The largest effect sizes were observed for plasma p-tau biomarkers, with plasma p-tau217 showing the most pronounced group differences (r ranging from 0.59 to 0.88, and percentage difference from 151 to 357%). Although statistically significant differences were observed for plasma Aβ42/40, the differences between biologically defined groups were overall smaller than those of plasma tau biomarkers. The same analysis was repeated using the Elecsys CSF p-tau181/Aβ42 to classify patients into AD and non-AD CSF profile, in the subset of patients where this measurement was available (Table S3; n=149) and the results were similar (Table S4). For most of the assays, the measurement in CSF was also available and the differences between groups are shown in Table S5.

**FIGURE 1.**
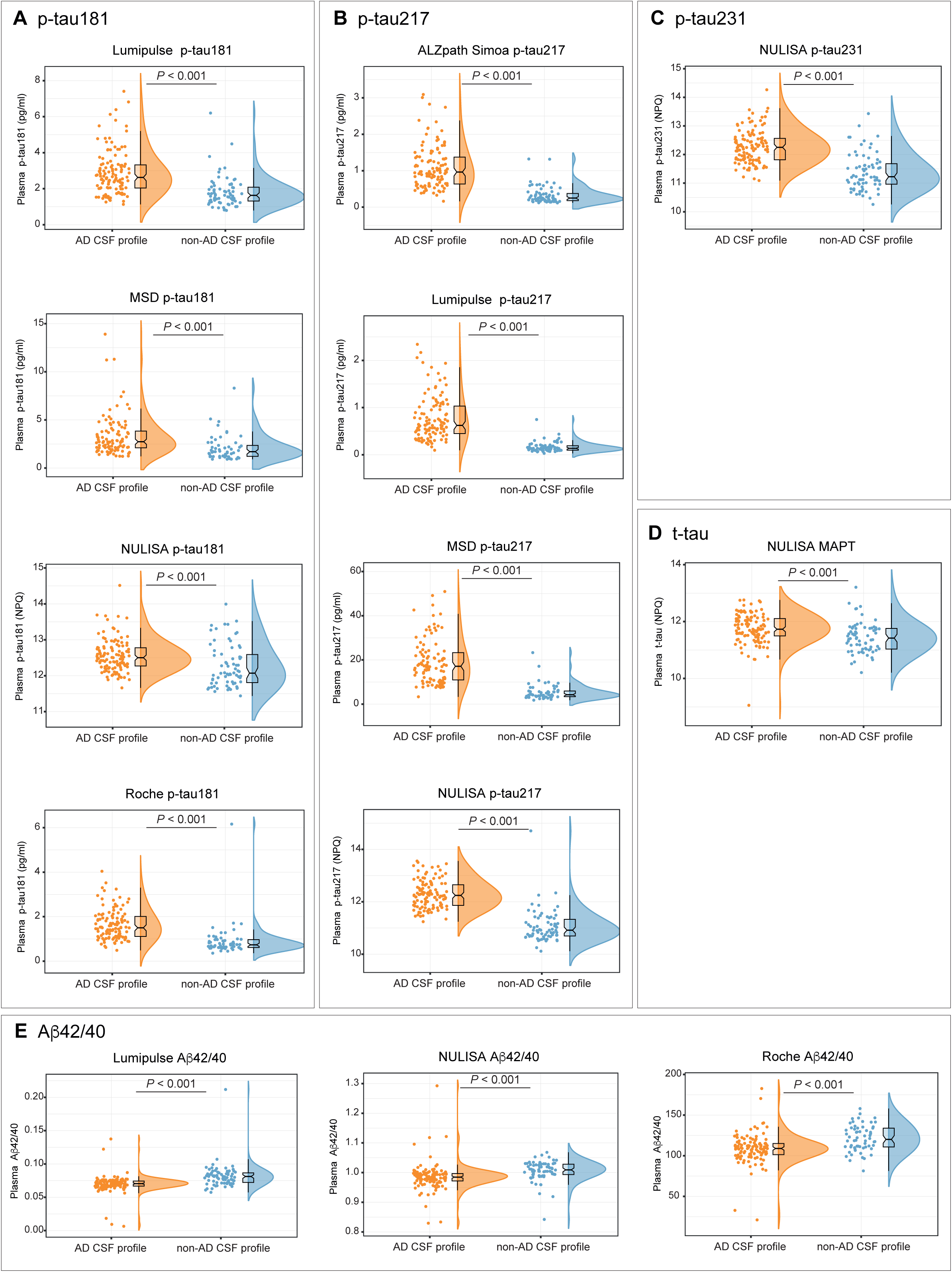
Raincloud plots showing differences in plasma biomarkers (A, p-tau181; B, p-tau217; C, p-tau231; D, t-tau; E, Aβ42/40) between the AD CSF profile (orange) and non-AD CSF profile (blue) groups. The box plot displays the median (horizontal line), interquartile range (box), and 1.5x interquartile range (whiskers). Individual biomarker values are also represented. Group differences were assessed using the Mann-Whitney *U* test. Table 2 presents the median and interquartile ranges for each group, along with the effect sizes of the differences.

### 3.3 Discrimination of AD CSF biomarker profile status

We next assessed the performance of plasma biomarkers to discriminate between AD and non-AD CSF profiles in a ROC curve analysis (Figure 2). The AUCs were as follows (from highest to lowest): Lumipulse p-tau217 (0.97, 95% CI 0.94-0.99), MSD p-tau217 (0.95, 95% CI 0.92-0.99), NULISA p-tau217 (0.95, CI 95% 0.91-0.99), ALZpath Simoa p-tau217 (0.94, 95% CI 0.90-0.98), Roche p-tau181 (0.89, 95% CI 0.84-0.94), NULISA p-tau231 (0.85, 95% CI 0.79-0.91), Lumipulse p-tau181 (0.81, 95% CI 0.74-0.87), Lumipulse Aβ42/40 (0.79, 95% CI 0.72-0.87), NULISA Aβ42/40 (0.77, 95% CI 0.69-0.84), MSD p-tau181 (0.75, 95% CI 0.67-0.84), Roche Aβ42/40 (0.73, 95% CI 0.64-0.81), NULISA MAPT (0.69, 95% CI 0.61-0.77) and NULISA p-tau181 (0.69, 95% CI 0.60-0.78). Sensitivities and Specificities of each assay are depicted in Table S6 and Table S7. After setting the sensitivity to 90%, all plasma p-tau217 biomarkers investigated had a specificity higher than 75%. ROC curve analyses were repeated using the Elecsys CSF p-tau181/Aβ42 to classify patients and the results were similar (Table S8).

**FIGURE 2.**
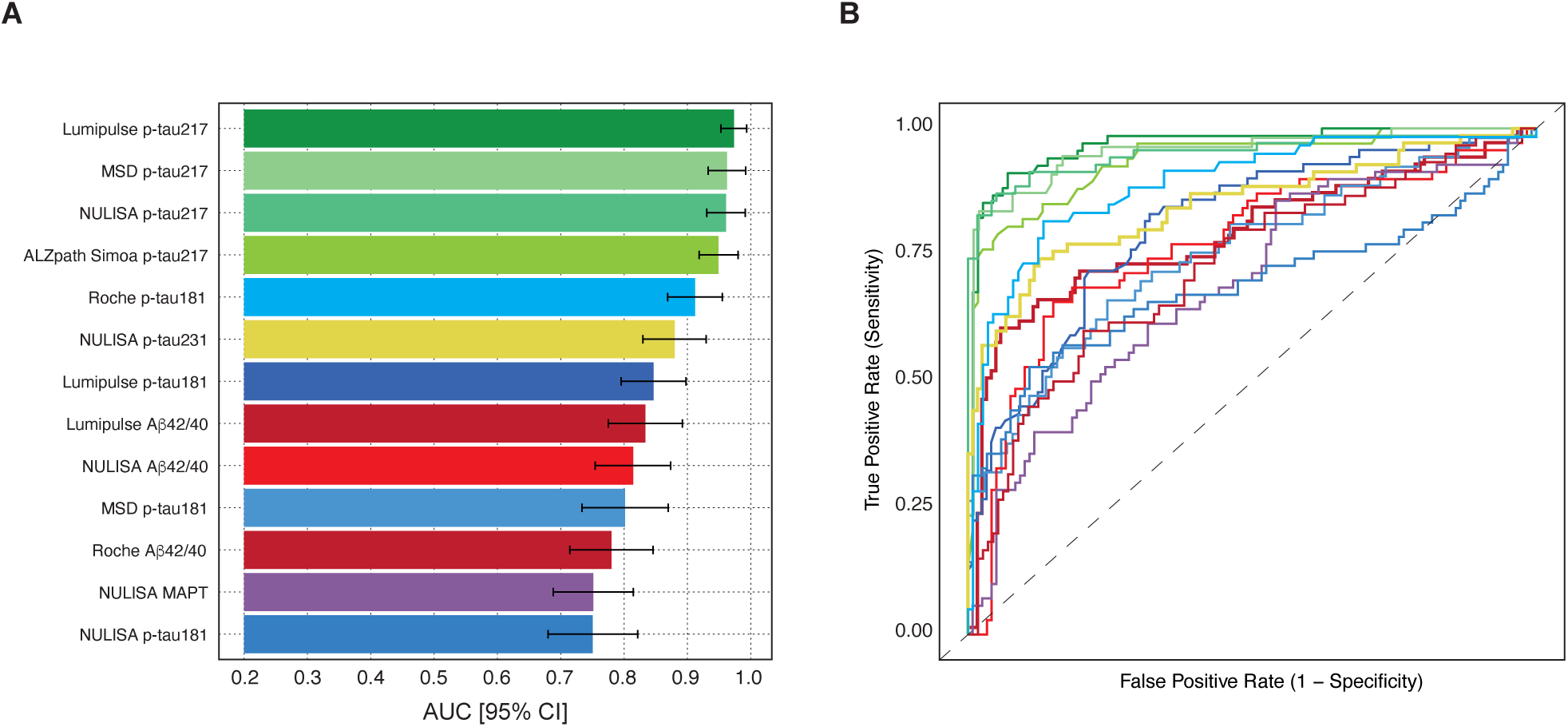
Diagnostic performance of plasma biomarkers in discriminating AD CSF profiles from non-AD CSF profiles using Receiver Operating Characteristic (ROC) analyses. (A) Bar plots showing the Areas Under the Curve (AUC) with their corresponding 95% confidence intervals (CIs) for each plasma biomarker. (B) ROC curves for the different plasma biomarkers, distinguished by color: p-tau181 (blue), p-tau217 (green), p-tau231 (yellow), t-tau (purple), and Aβ42/40 (red).

### 3.4 Sensitivity analyses

To account for missing plasma biomarker data in some patients and to ensure that variations in participant numbers across different assays did not affect the results, we performed a sensitivity analysis on the subgroup of patients for whom all plasma biomarkers were available (Table S9). This analysis also included the plasma biomarkers previously reported by our group,^12^ enabling a direct comparison of 22 plasma biomarkers across 141 patients. The analysis comprised 17 plasma p-tau assays (8 p-tau181, 6 p-tau217, 3 p-tau231), 2 t-tau assays, and 3 Aβ42/40 assays (Figure S1 and Table S10). Remarkably, among all the plasma biomarkers, all six plasma p-tau217 assays achieved an AUC above 0.90. Additionally, two plasma p-tau181 assays also had an AUC exceeding 0.90 (Figure S1).

Given the inclusion of a range of different clinical diagnoses in the study (reflecting the reality of a memory clinic), we conducted additional sensitivity analyses focused on patients with the syndromic diagnoses of SCD, MCI, and AD dementia (Table S11). To evaluate plasma biomarkers in the early stages, we also performed an analysis including only patients with SCD and MCI (Table S12). The AUC results from these sensitivity analyses (Table S13) were consistent with those obtained from the full sample.

### 3.5. Correlations between biomarkers

Finally, we assessed the correlation between plasma biomarkers and their corresponding CSF counterparts across the assays where both measures were available (Lumipulse p-tau181, MSD p-tau181, NULISA p-tau181, Roche p-tau181, Lumipulse p-tau217, MSD p-tau217, NULISA p-tau217, NULISA p-tau231, and NULISA MAPT; Figure S2). All correlations were statistically significant, with the highest Spearman coefficients observed for the p-tau217 assays (Lumipulse p-tau217: r=0.81; NULISA p-tau217: r=0.73; MSD p-tau217: r=0.78). Additionally, a matrix of plasma biomarker correlations is included in Figure S3.

## 4. DISCUSSION

In this study, we conducted a head-to-head comparison of several plasma tau and Aβ42/40 biomarkers in a clinical cohort from a memory unit, encompassing patients with a wide range of cognitive and/or behavioural symptoms. We found that several plasma biomarkers exhibited high performance in detecting AD, though notable differences among them are worth highlighting. First, plasma p-tau biomarkers generally demonstrated higher performance in classifying Aβ status and showed a larger effect size between groups compared to plasma total tau and Aβ42/40 assays. Second, all plasma p-tau217 assays investigated showed excellent performance, often surpassing those assays targeting other phosphorylation sites, although some assays targeting T181 and T231 also produced convincing results. These findings were consistent in sensitivity analyses across different syndromic diagnoses, reflecting routine clinical practice. Finally, p-tau217 assays showed the highest correlation between plasma and CSF levels, suggesting that this plasma biomarker more accurately reflects CSF levels. Notably, plasma p-tau217 levels in the non-AD group are generally low, allowing for clearer differentiation from the AD group.

Building on our previous head-to-head study,^12^ we now provide a direct comparison of 22 plasma biomarkers, including 8 p-tau181, 6 p-tau217, 3 p-tau231, 2 t-tau assays, and 3 Aβ42/40 assays. This direct comparison within the same cohort, and in a subset of identical patients, allows for a precise evaluation of these assays under consistent conditions. The promising outcome is that several assays, particularly all p-tau217 assays and some p-tau181 assays, exhibit high performance to classify Aβ status (with AUCs above 90%), making them potentially well-suited for clinical practice (contingent upon regulatory approval). Notably, all plasma p-tau217 assays meet the recently established minimum performance criteria for blood biomarkers set by the BBM Workgroup, which recommend a sensitivity of 90% and a specificity above 75% or higher for use as a triaging test in specialized centers.^24^ These results are consistent with previous reports indicating that plasma p-tau217 assays have the best performance in symptomatic patients.^4,6,9–13^

Although statistically significant differences were observed for plasma Aβ42/40 biomarkers, the effect sizes were small (r<0.30). This aligns with previous research showing only modest reductions (8-15%) in blood Aβ levels in AD.^25^ The small effect sizes in blood Aβ levels may impact the robustness of the analysis, as minor variations can influence patient classification.^26,27^

Our study evaluates a diverse range of blood-based biomarkers, covering various phosphorylation sites, methods, and platforms. The results show high diagnostic performance for several assays, underscoring an expanding array of options for AD diagnosis. While our study focused on immunoassays and did not include IP-MS technologies —despite their excellent performance but limited suitability for widespread clinical use— we did incorporate the novel NULISA platform.^22^ This platform, based on proximity ligation assay technology and capable of multiplexing, demonstrated comparable performance to some of the top immunoassays. Ultimately, the choice of biomarker may be influenced by factors such as context, availability, cost, regulatory status, and scalability, with some assays being fully automated and others semi-automated.

This study has several limitations. First, it lacks pathological confirmation of AD, which is still considered as the gold standard, or amyloid/tau PET imaging. Second, it is unicentric, meaning the findings are based on patients from our hospital and need validation in other cohorts globally. Third, the study did not explore potential confounding factors, clinical robustness, or assess the clinical impact of using these plasma biomarkers. However, the study’s strengths include its focus on a population referred from primary care for cognitive and behavioural assessment at a public hospital in Barcelona, which serves as a reference hospital for the area. This population is more heterogeneous, with a greater range of clinical presentations compared to only research cohorts. Moreover, we compared the performance of blood-based biomarkers with the current routine diagnostic test, CSF analysis, providing a realistic reflection of clinical practice.

In conclusion, our study shows that several plasma biomarkers, particularly p-tau217 and some p-tau181 assays, have high performance in detecting AD pathology in a memory clinic setting. These findings support the implementation of these tests in clinical practice.

## Data Availability

All requests for raw and analysed data and materials will be promptly reviewed by the senior authors to verify whether the request is subject to any intellectual property or confidentiality obligations. Bulk Anonymized data can be shared by request from any qualified investigator for the sole purpose of replicating procedures and results presented in the article, providing data transfer agrees with EU legislation and decisions by the IRB of each participating center.

## ACKNOWLEDGMENTS

The authors would like to express their most sincere gratitude to the BIODEGMAR participants and relatives without whom this research would have not been possible. The authors thank to Helena Blasco, Marina de Diego, Juan José Hernández Sánchez, and Esther Jiménez-Moyano and all the staff of Barcelonaβeta Brain Research Center, the Neurology Department of Hospital del Mar and the Laboratori de Referència de Catalunya for technical support. We thank Theresa A. Day, Jeroen Vanbrabant, Erik Stoops, Eugeen Vanmechelen, Gallen Triana-Baltzer, Setareh Moughadam, Hartmuth Kolb, and Jeff L. Dage for previously providing biomarkers measurements in the BIODEGMAR cohort and critically reviewing the manuscript.

## CONFLICTS OF INTEREST

AS, QH and BZ are employees of Alamar Biosciencies, Inc. NLB and AN are employees of Fujirebio Europe N.V. and Fujirebio Iberia, respectively. TM is an employee of Meso Scale Diagnostics, LLC. GK is a full-time employee of Roche Diagnostics GmbH. MC is a full-time employee of Roche Diagnostics International Ltd and an owner of shares in Roche. HZ has served at scientific advisory boards and/or as a consultant for Abbvie, Acumen, Alector, Alzinova, ALZPath, Amylyx, Annexon, Apellis, Artery Therapeutics, AZTherapies, Cognito Therapeutics, CogRx, Denali, Eisai, LabCorp, Merry Life, Nervgen, Novo Nordisk, Optoceutics, Passage Bio, Pinteon Therapeutics, Prothena, Red Abbey Labs, reMYND, Roche, Samumed, Siemens Healthineers, Triplet Therapeutics, and Wave, has given lectures sponsored by Alzecure, BioArctic, Biogen, Cellectricon, Fujirebio, Lilly, Novo Nordisk, Roche, and WebMD, and is a co-founder of Brain Biomarker Solutions in Gothenburg AB (BBS), which is a part of the GU Ventures Incubator Program (outside submitted work). AP-P has served on advisory boards for Schwabe Farma Iberica. MS-C has received in the past 36mo consultancy/speaker fees (paid to the institution) from by Almirall, Eli Lilly, Novo Nordisk, and Roche Diagnostics. He has received consultancy fees or served on advisory boards (paid to the institution) of Eli Lilly, Grifols and Roche Diagnostics. He was granted a project and is a site investigator of a clinical trial (funded to the institution) by Roche Diagnostics. In-kind support for research (to the institution) was received from ADx Neurosciences, Alamar Biosciences, ALZPath, Avid Radiopharmaceuticals, Eli Lilly, Fujirebio, Janssen Research & Development, Meso Scale Discovery, and Roche Diagnostics; MS-C did not receive any personal compensation from these organizations or any other for-profit organization.

## FUNDING SOURCES

FA receives funding from the JDC2022-049347-I grant, funded by the MCIU/AEI/10.13039/501100011033 and the European Union NextGenerationEU/PRTR. HZ is a Wallenberg Scholar and a Distinguished Professor at the Swedish Research Council supported by grants from the Swedish Research Council (#2023-00356; #2022-01018 and #2019-02397), the European Union’s Horizon Europe research and innovation programme under grant agreement No 101053962, Swedish State Support for Clinical Research (#ALFGBG-71320), the Alzheimer Drug Discovery Foundation (ADDF), USA (#201809-2016862), the AD Strategic Fund and the Alzheimer’s Association (#ADSF-21-831376-C, #ADSF-21-831381-C, #ADSF-21-831377-C, and #ADSF-24-1284328-C), the European Partnership on Metrology, co-financed from the European Union’s Horizon Europe Research and Innovation Programme and by the Participating States (NEuroBioStand, #22HLT07), the Bluefield Project, Cure Alzheimer’s Fund, the Olav Thon Foundation, the Erling-Persson Family Foundation, Familjen Rönströms Stiftelse, Stiftelsen för Gamla Tjänarinnor, Hjärnfonden, Sweden (#FO2022-0270), the European Union’s Horizon 2020 research and innovation programme under the Marie Skłodowska-Curie grant agreement No 860197 (MIRIADE), the European Union Joint Programme – Neurodegenerative Disease Research (JPND2021-00694), the National Institute for Health and Care Research University College London Hospitals Biomedical Research Centre, and the UK Dementia Research Institute at UCL (UKDRI-1003). MS-C receives funding from the European Research Council (ERC) under the European Union’s Horizon 2020 research and innovation programme (Grant agreement No. 948677); ERA PerMed (ERAPERMED2021-184); Project “PI19/00155” and “PI22/00456, funded by Instituto de Salud Carlos III (ISCIII) and co-funded by the European Union; and from a fellowship from ”la Caixa” Foundation (ID 100010434) and from the European Union’s Horizon 2020 research and innovation programme under the Marie Skłodowska-Curie grant agreement No 847648 (LCF/BQ/PR21/11840004).

ELECSYS is a trademark of Roche. All other product names and trademarks are the property of their respective owners. Elecsys β-amyloid(1–42) CSF and Elecsys Phospho-Tau (181P) CSF assays are approved for clinical use. The NeuroToolKit is a panel of exploratory prototype assays designed to robustly evaluate biomarkers associated with key pathologic events characteristic of AD and other neurological disorders, used for research purposes only and not approved for clinical use (Roche Diagnostics International Ltd, Rotkreuz, Switzerland).

Alamar, ALZpath, Fujirebio, MSD and Roche Diagnostics provided reagents and/or biomarker measurements in-kind. A few employees of these companies, listed as co-authors, made direct contributions to this research (see the Contributors section). The companies were not involved in the study design, or the analysis and interpretation of the data. The corresponding authors had full access to all the data in the study and had final responsibility for the decision to submit for publication.

## ETHICS APPROVAL AND CONSENT TO PARTICIPATE

The BIODEGMAR study was approved by the Independent Ethics Committee “Parc de Salut Mar”, Barcelona (CEIC PSMAR, project code 2018/7805I). All participants from BIODEGMAR provided informed consent.

## DATA SHARING STATEMENT

Not applicable.

## AUTHOR CONTRIBUTIONS

Federica Anastasi and Aida Fernández-Lebrero are co–first authors. Albert Puig-Pijoan, and Marc Suárez-Calvet are co–senior authors. Albert Puig-Pijoan and Marc Suárez-Calvet had full access to all the data in the study and take responsibility for the integrity of the data and the accuracy of the data analysis.

### Concept and design

Nicholas J. Ashton and Marc Suárez-Calvet.

### Acquisition, analysis, or interpretation of data

Federica Anastasi, Aida Fernández-Lebrero, Nicholas J. Ashton, Paula Ortiz-Romero, Javier Torres-Torronteras, Armand González-Escalante, Marta Milà-Alomà, José Contador, Greta García-Escobar, Rosa-María Manero-Borràs, Irene Navalpotro-Gómez, Aparna Sahajan, Qinyu Hao, Bingqing Zhang, Andreas Jeromin, Nathalie Le Bastard, Alicia Nadal, Tahmine Mousavi, Gwendlyn Kollmorgen, Margherita Carboni, Oriol Grau-Rivera, Marta del Campo, Henrik Zetterberg, Kaj Blennow, Albert Puig-Pijoan, and Marc Suárez-Calvet.

### Drafting of the manuscript

Federica Anastasi, Aida Fernández-Lebrero, Albert Puig-Pijoan, Marc Suárez-Calvet.

### Statistical analysis

Federica Anastasi, Aida Fernández-Lebrero, Armand González-Escalante, Marta Milà-Alomà, Albert Puig-Pijoan, Marc Suárez-Calvet.

### Obtained funding

Henrik Zetterberg, Kaj Blennow, Marc Suárez-Calvet.

### Administrative, technical, or material support

Aida Fernández-Lebrero, Greta García-Escobar, Paula Ortiz-Romero, Aparna Sahajan, Qinyu Hao, Bingqing Zhang, Andreas Jeromin, Nathalie Le Bastard, Alicia Nadal, Tahmine Mousavi, Gwendlyn Kollmorgen, Margherita Carboni.

### Supervision

Albert Puig-Pijoan, Marc Suárez-Calvet.

*All authors critically reviewed and approved the final manuscript*.

---

**Figure S1.**
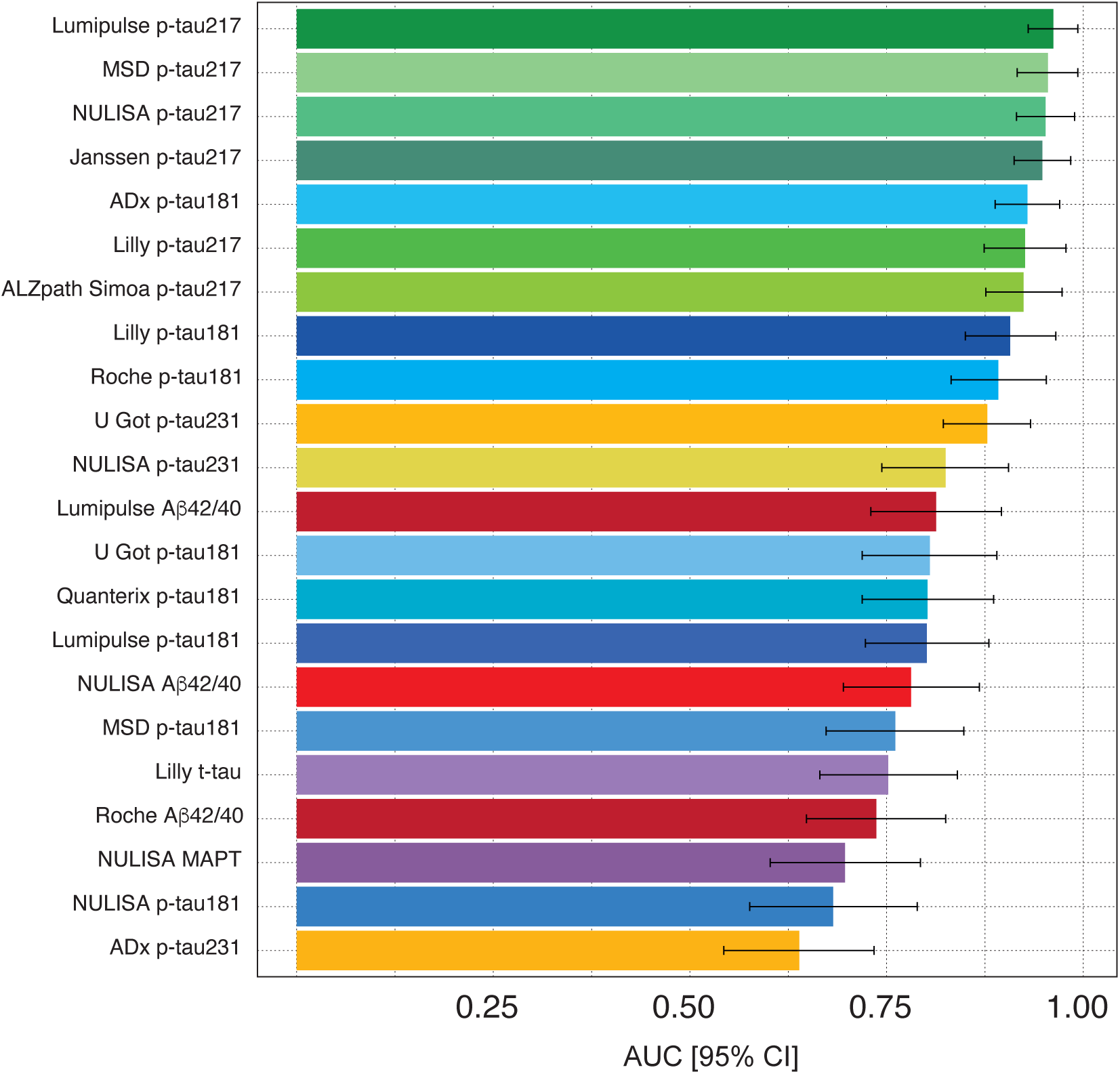

**Figure S2.**
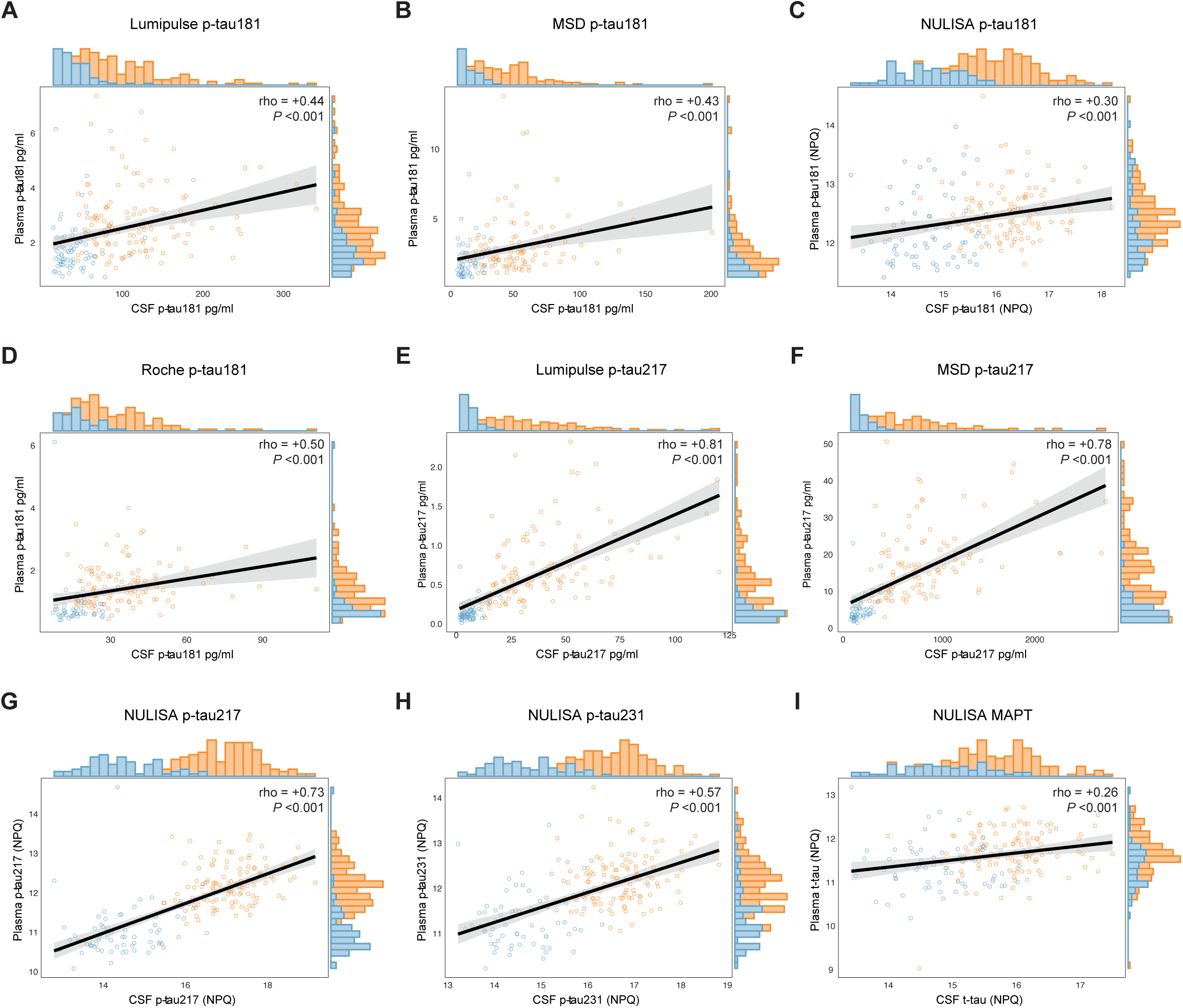

**Figure S3.**
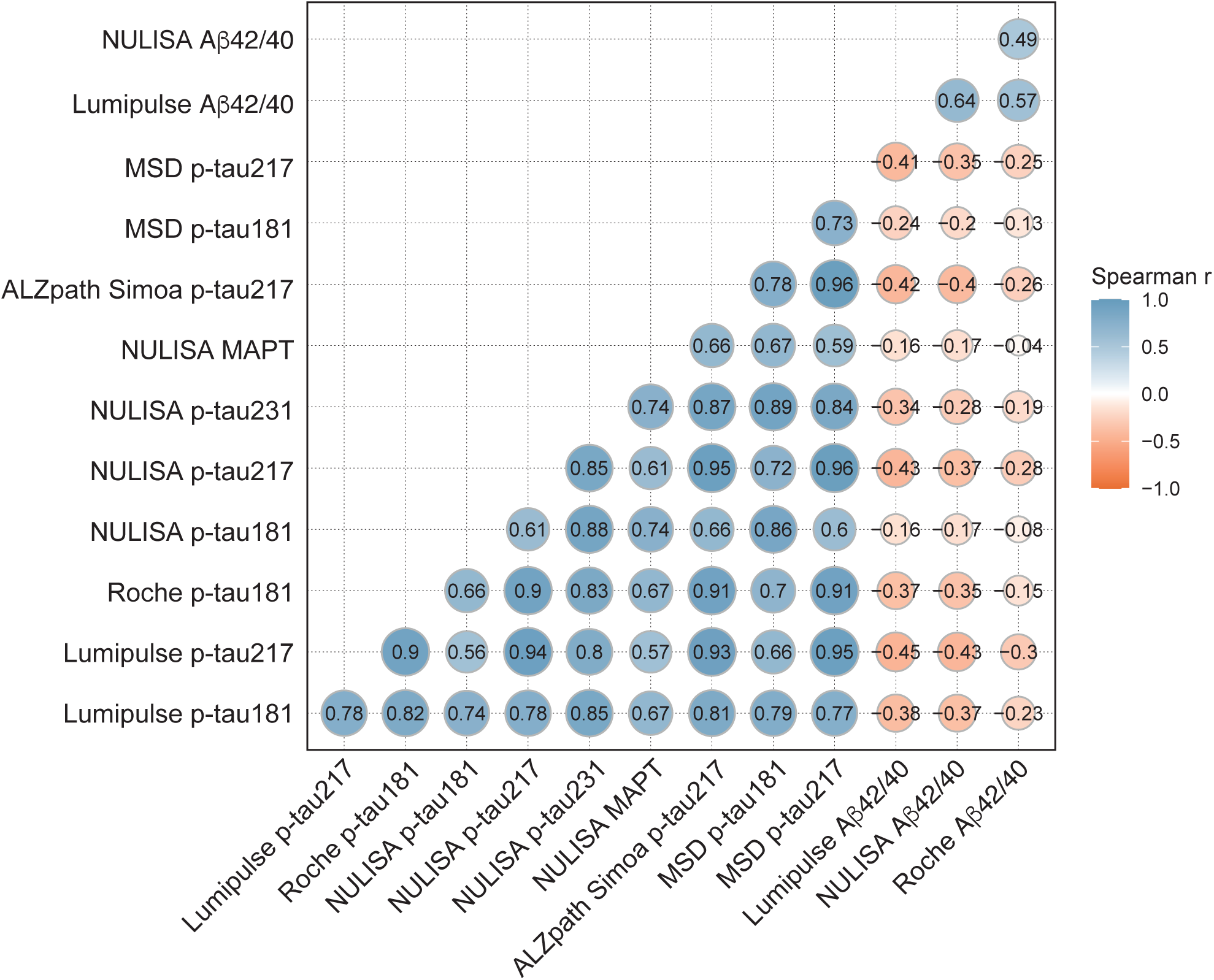

## Notes

### Author Declarations

The BIODEGMAR study was approved by the Independent Ethics Committee Parc de Salut Mar, Barcelona (CEIC PSMAR, project code 2018/7805I). All participants from BIODEGMAR provided informed consent.

